# Missed diagnoses of giant cell arteritis in acute ocular ischemia: A diagnostic blind spot

**DOI:** 10.1101/2025.05.20.25328006

**Authors:** Jagannadha Avasarala, Suhas Gangadhara, Joseph Bobadilla, Romil Chadha

## Abstract

**Background:** In patients >50 years of age, isolated acute visual symptoms may represent occult giant cell arteritis (GCA). Additionally, central retinal artery occlusion (CRAO) or ischemic optic neuropathy (ION) may present with isolated, acute ophthalmic symptoms. In such settings, GCA, a treatable vasculitis, is under-diagnosed or misdiagnosed, with catastrophic consequences.

**Objective:** To quantify diagnostic gaps in GCA evaluation among patients presenting with ocular ischemic symptoms in emergency settings, using both institutional and national datasets.

**Methods:** We conducted a retrospective cohort analysis using data from patients aged >50 years presenting to the emergency department (ED) with isolated ocular symptoms between 2021 and 2024. Datasets included the University of Kentucky (UKY) Epic EHR and Epic Cosmos (299 million patients nationwide). Patients were stratified into three cohorts: occult GCA, CRAO, and ION. Multivariable logistic regression was performed using standard statistical software to estimate predictors of GCA diagnosis, adjusting for temporal artery ultrasound (TAUS), temporal artery biopsy (TAB) and use of corticosteroid administration prior to a diagnosis.

**Results:** In the UKY dataset of 103 patients with acute ocular symptoms, 38.8% had confirmed GCA, 27.2% were clinically diagnosed, and 34% were ruled out. Among confirmed cases, 67.5% underwent TAUS and 17.5% had a TAB. A total of 66/103 (64%) received steroids before testing and 46/66 (69.7%) of them tested negative for GCA. In Epic Cosmos (nationwide) data, 62.8% of 17,372 patients lacked any testing; TAUS-specific data were unavailable.

Among patients with CRAO (n=90) in the UKY cohort, 12.2% underwent TAUS, with 63.6% diagnosed with GCA; workup for stroke revealed an embolic stroke 22/90 (24.4%); 75.5% received no further workup after stroke evaluation was unrevealing and 1/90 underwent TAB that was negative, the patient received steroids prior. In Epic Cosmos, of 18,621 patients with CRAO, 561 underwent TAB (3%), and 5908 (31%) had ultrasound but TAUS-specific data was unavailable.

Among 594 UKY patients with ischemic optic neuropathy (ION), 28 had carotid ultrasound or TAUS; 15 had TAUS, with 33% diagnosed as having GCA; 9 underwent TAB and 33% had GCA. Additionally, 94.1% of the UKY cohort had no testing for GCA; In Epic Cosmos, of the 56554 patients with ION, 2686 (4.7%) underwent carotid U/S or TAUS, or both. 901(1.6%) underwent TAB, while 52967/56554 (93.6%) had no formal testing for GCA.

**Conclusions:** GCA is underdiagnosed among patients presenting with acute ocular ischemic syndromes and TAUS, a point-of-care non-invasive diagnostic tool, is underutilized. Integrating TAUS into ED protocols for ocular ischemia could reduce diagnostic delays and prevent irreversible blindness.

## Introduction

The most common systemic, autoimmune, treatable, large-vessel vasculitis in adults ≥50 years of age is GCA. It is a leading cause of permanent vision loss^1^ if not promptly diagnosed or left untreated. Diagnostic delays significantly increase the risk of visual loss^2^ and ischemic complications^3^. Incidence rates of GCA vary from 18 to 29 per 100,000 individuals ≥ 50 years of age, with higher rates in European populations, about 41–113 per 100,000^4^. Although stroke is a rare complication of GCA, it predominantly involves the vertebrobasilar circulation in the elderly^5^. Despite decades of clinical awareness, diagnostic delays persist, especially when GCA presents without systemic symptoms, the occult GCA variant, underscoring that GCA could be a spectrum disorder. Patients ≥ 50 years of age presenting with new ocular symptoms restricted to the eye may harbor occult GCA in up to 20–38% of cases^6–9^. A wide variety of ocular manifestations—including amaurosis fugax, diplopia, fluctuating vision, ptosis, anisocoria, cranial nerve palsies, and internuclear ophthalmoplegia—should prompt suspicion of GCA^10^.

Acute ocular ischemia, including CRAO and ION, may be the first or only manifestation of GCA. The American Heart Association (AHA) recognizes CRAO as a stroke since retinal ischemia is caused by CRAO; additionally, an arteritic CRAO is described^11^. Moreover, CRAO and cardiovascular risk factors and carotid artery disease^12^ can be linked. The American Academy of Ophthalmology (AAO) defines arteritic ION as an ophthalmologic emergency requiring immediate recognition and intervention^13^. Arteritic ION, which can result from GCA, necessitates urgent diagnosis to prevent permanent vision loss^14^. In such cases, diagnosis hinges on early suspicion and appropriate vascular imaging, particularly TAUS, particularly if a real-time diagnosis is sought. However, diagnostic pathways remain fragmented and while the workup of CRAO is often restricted to primary stroke syndrome with minimal investigations for GCA, ION may be attributed to a diagnosis non-arteritic ION (NAION) without workup for an arteritic ION.

The broader implications of delayed or missed GCA diagnosis are profound. By 2050, GCA-related blindness is projected to affect half a million individuals globally^15^ with associated U.S. healthcare costs exceeding $ 77 billion in visual impairment and $ 6.6 billion in direct treatment expenses^16^. Despite high stakes, no formal guidelines currently exist to use TAUS as a frontline diagnostic tool for occult GCA presentations. Although the 2023 European Alliance of Associations for Rheumatology (EULAR) recommendations endorse imaging modalities such as TAUS for GCA diagnosis^17^, they do not explicitly address isolated ocular presentations presenting without systemic symptoms.

The 2023 EULAR recommendations endorse imaging as first-line for large vessel vasculitis, but its use in the emergency department (ED), remains limited. In this study, we analyzed both institutional and nationwide data to determine how often GCA is evaluated in patients presenting with isolated ocular symptoms and whether TAUS and TAB are appropriately utilized.

## Methods

We conducted a retrospective cohort study approved by the UKY IRB in accordance with the Declaration of Helsinki and HIPAA regulations. Deidentified patient data were extracted from two sources: (1) the UKY Epic electronic health record (EHR) system, and (2) Epic Cosmos, a national, federated dataset comprising over 299 million patients. The study period spanned from January 2021 to December 2024, and included patients aged >50 years who presented to the emergency department (ED) with acute ocular symptoms.

Patients were categorized into three clinical cohorts: (1) isolated ocular symptoms concerning for occult GCA, (2) CRAO, and (3) ION). Inclusion criteria required documentation of acute visual changes and/or a diagnostic label of CRAO or ION. Diagnostic evaluations of interest included TAUS, carotid duplex ultrasound, TAB, stroke imaging (CT/MRI), and the use of corticosteroids initiated prior to testing.

Data for TAUS was extracted directly from the institutional EHR. In Epic Cosmos, procedural codes for carotid ultrasound and TAUS are not separable due to shared CPT coding; as a result, TAUS-specific diagnostic yield cannot be calculated from this national dataset. Likewise, individual-level TAB outcomes were not available in Cosmos and were excluded from regression analysis.

For the UKY cohorts, odds ratios (ORs) and 95% confidence intervals (CIs) were derived using multivariable logistic regression with GCA diagnosis (biopsy-proven or clinically adjudicated) as the binary outcome. Independent variables included TAUS performance, TAB performance, and corticosteroid use prior to diagnostic testing. Modeling was implemented in Python using statsmodels, and the logistic regression model was specified as,

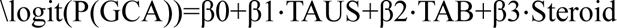

Where regression modeling was infeasible due to low event counts (e.g., in CRAO and ION subgroups), ORs and CIs were estimated from raw proportions. For the Epic Cosmos dataset, only descriptive statistics on test utilization were reported due to the lack of patient-level outcome linkage. A comparative forest plot, ROC curve (AUC), and annotated Python code were generated to illustrate diagnostic performance across TAUS, TAB, and steroid use in patients presenting with occult GCA, CRAO, or ION. Analyses were conducted using Python (statsmodels, scikit-learn, matplotlib), and all code is archived for reproducibility. All statistical analyses were independently verified, and the code was reviewed for accuracy and reproducibility by a trained professional in data modeling, coding and analysis.

## Statistical Analysis

We performed a multivariable logistic regression analysis using patient-level data from the full University of Kentucky cohort (n=787), which included patients presenting with occult GCA, CRAO and ION. The binary outcome was GCA diagnosis (confirmed or clinically diagnosed vs. ruled out). Predictor variables included performance of TAUS, TAB, and corticosteroid administration prior to diagnostic testing. Odds ratios with 95% confidence intervals were derived using maximum likelihood estimation. Model discrimination was assessed using a receiver operating characteristic (ROC) curve, and diagnostic performance was visualized using a forest plot. Area under the curve (AUC) was 0.71. Analyses were conducted using Python (statsmodels, scikit-learn, matplotlib), and all code is archived for reproducibility. In general, TAUS was the strongest predictor of GCA diagnosis, while steroid use prior to testing diminished diagnostic sensitivity.

## Results

Among the 103 with isolated acute ocular symptoms (occult GCA) in the UKY dataset 40 (38.8%) had a confirmed diagnosis of GCA, 28 (27.2%) were clinically diagnosed, and 35 (34%) ruled out. Of those confirmed, 27 (67.5%) had TAUS, and 7 (17.5%), temporal artery biopsy (TAB). Additionally, 6 (15%) underwent confirmation with both TAUS and TAB. A total of 66/103 (64%) received steroids before testing and 46/66 (69.7%) of them tested negative for GCA. In Epic Cosmos data, 62.8% of 17,372 patients lacked any testing; TAUS-specific data were unavailable.

90 patients underwent workup for CRAO in the UKY dataset; 11 (12.2%) underwent TAUS and 7/11 (63.6%) had GCA; 22/90 (24.4%) had an embolic source; 68/90 (75.5%) had no workup after stroke evaluation was unrevealing and 1/90 underwent TAB, post-steroids that was negative. In Epic Cosmos, of 18,621 patients with CRAO, 561 underwent TAB (3%), and 5908 (31%) had ultrasound but TAUS-specific data was unavailable.

Of 594 patients with ION in the UKY dataset, 28/594 (4.7%) underwent carotid U/S or TAUS, 15/28 (53.5%) underwent TAUS, 5/15 (33%) had GCA; 9/594 underwent TAB, and 3/9 (33%) had GCA. Of the rest, 559/594 (94.1%) did not have any testing for GCA. In Epic Cosmos, 56554 patients with ION, 2686 (4.7%) underwent carotid U/S or TAUS, or both. 901(1.6%) underwent TAB, while 52967/56554 (93.6%) had no formal testing for GCA or stroke.

Multivariable logistic regression modeling was performed on the full UKY cohort (n=787; combining patients with occult GCA, CRAO, and ION). It demonstrated that TAUS was the strongest independent predictor of a GCA diagnosis (OR 12.74; 95% CI 7.04–23.05) and the only testing modality with a point-of-care utility. Next, TAB was also significantly associated with GCA (OR 4.68; 95% CI 1.57–13.99), albeit not point-of-care. Prior corticosteroid administration yielded some association (OR 1.74; 95% CI 1.03–2.95), likely reflecting selection bias toward sicker patients. Our findings validate TAUS as a powerful, real-time diagnostic modality highlighting the need for imaging protocols prior to steroid initiation. The model’s discrimination was robust, with an AUC of 0.71 further underscoring the diagnostic utility of TAUS in acute ocular presentations of GCA.

In the Epic Cosmos dataset, 62.8% of patients with presumed occult GCA (n=17,372) had no diagnostic testing—neither temporal artery ultrasound (TAUS) nor biopsy—raising the risk of missed diagnoses with potentially catastrophic consequences. Among patients with ION, 93-94% had no formal GCA workup in either UKY or Cosmos datasets. Similarly, 66% of CRAO patients in Cosmos lacked any diagnostic evaluation. Across all subgroups (occult GCA, CRAO, ION), 62–94% of patients in Cosmos received no GCA testing.

## Discussion

This study reveals a persistent diagnostic void in the emergency evaluation of ocular ischemic events in patients over 50 years of age. Specifically, GCA is a treatable but often unrecognized disease and our findings show that in institutional and national datasets, over 90% of patients with CRAO, ION, and 62.8% of patients with isolated visual symptoms in the Cosmos dataset had no evaluation for GCA. The AHA recognizes CRAO as a stroke^11^ and the AAO notes that ION is an ophthalmological emergency^13^. The fact that the majority of patients with symptoms worrisome for GCA remain unscreened presents an opportunity for a change. To our knowledge, no prior study has interrogated acute, isolated ocular presentations—occult GCA, CRAO, or ION—in emergency settings with a focus on GCA after mimics were excluded.

In our study, TAUS emerged as the strongest independent predictor of GCA diagnosis, outperforming TAB in sensitivity while maintaining a practical edge in speed and safety. Multivariable analysis of the full UKY cohort (n=787) showed that TAUS was the strongest predictor of GCA (OR 12.74; 95% CI 7.04–23.05), and the only diagnostic tool with true point-of-care utility. Next, TAB showed a significant association (OR 4.68; 95% CI 1.57–13.99), but lacks immediate applicability. Our results emphasize the value of TAUS in real-time diagnosis and underscore the importance of imaging before steroid exposure.

The underuse of TAUS, particularly in EDs, represents a critical gap in care. Corticosteroid initiation prior to diagnostic workup significantly reduced test sensitivity, underscoring the importance of early imaging. Our findings support the urgent integration of TAUS into emergency stroke protocols and ophthalmic triage pathways. Patients presenting with acute vision loss should be screened for GCA, especially when conventional stroke evaluation findings are unrevealing. As well, TAB remains a valuable secondary tool, particularly when TAUS is inconclusive. The TABUL study^18^ provides robust evidence favoring TAUS over TAB. Sensitivity of TAB was only 39% compared to 54% for TAUS, while specificity favored TAB (100%) over TAUS (81%)^10^. In the TABUL study, a strategy of reserving TAB for negative TAUS cases increased diagnostic sensitivity to 65% and reduced the need for biopsies by 43%. These findings support the integration of TAUS as the first-line diagnostic tool in GCA.

A critical and under-recognized failure in clinical practice is the misdiagnosis of arteritic ischemic optic neuropathy (AION) or posterior ischemic optic neuropathy (PION)—both manifestations of GCA—as non-arteritic ION (NAION), purely due to failure to perform TAUS. In the absence of systemic symptoms, physicians frequently default to a NAION diagnosis without considering arteritic AION or PION. This results in a missed opportunity for life- and sight-saving steroid therapy. Unlike NAION, which has limited therapeutic options, arteritic AION and PION are both treatable with immediate corticosteroids. Our data show that in real-world settings, TAUS is either not ordered or misunderstood as a redundant test—despite its ability to identify occult or atypical GCA presentations in patients with acute ocular symptoms. This diagnostic inertia perpetuates irreversible blindness that could otherwise be prevented. TAUS should be a frontline test in all patients over 50 presenting with unexplained acute visual symptoms—particularly those presumptively labeled as NAION without confirmatory vascular imaging.

The 2022 American College of Rheumatology/European League Against Rheumatism (ACR/EULAR) classification criteria^19^ used a scoring system that can be adapted to screen patients in acute care settings, to exploit the diagnostic reach of TAUS and perhaps ED settings can utilize this concept.

Since there are an estimated 2–3 million annual ED visits for eye-related issues in the United States, and the projected shortage of ophthalmologists^20^ is a concern, frontline ED protocols must adapt. Public health initiatives akin to Get With The Guidelines® for stroke must be utilized to diagnose GCA in the context of acute ocular symptoms. Stroke education must be extended to include emergent ischemic vascular events involving the eye as acute ischemic ocular events are potential strokes.

## Study limitations

While the logistic regression model demonstrated high discriminative ability (AUC = 0.71) in the combined UKY cohort, this result should be interpreted with caution given the small sample size, single-center bias, and lack of external validation using national data. We realize that AUCs calculated from small samples are prone to overfitting, especially when the predictor (TAUS) is a strong discriminator in a single-center cohort with homogenous protocols. The performance of this model may not generalize to broader populations.

The UKY data likely had a higher TAUS utilization and better documentation, which inflates the model’s predictive accuracy. In Epic Cosmos, TAUS was poorly coded and could not be isolated from carotid Doppler tests, limiting external validation. We included patients who received diagnostic workups (TAUS), that causes selection and ascertainment bias may exclude patients who were missed entirely, and the dataset is already enriched for diagnosable GCA cases. Although our findings are subject to ascertainment bias since TAUS use predicts GCA we contend this reflects the clinical reality: without TAUS, GCA remains invisible. The absence of testing is not a statistical artifact but a systems-level failure.

Our study underscores the urgency of implementing TAUS in ED pathways for patients over 50 years of age with acute visual symptoms, where current practice may lead to diagnostic omission and irreversible blindness. In the real world, ED physicians, ophthalmologists and neurologists must often act on incomplete clinical data, and quickly, to establish a diagnosis that is treatable. Our study shows that TAUS is not only a frontline diagnostic tool that can help diagnose GCA, it is the only solution to establish a diagnosis in real time.

In our findings, GCA diagnosis included both biopsy-proven and clinically adjudicated cases. This may introduce subjectivity and heterogeneity in the outcome definition, affecting model stability. The model has not been tested on an independent cohort. Without external validation, the high AUC may reflect internal coherence but not real-world performance.

## Data Availability

All Data produced in the present study are available upon reasonable request to the authors.

